# Rescaling and Small Area Estimation of Health Survey Data as applied to Smoking Rates in Allegheny County, Pennsylvania

**DOI:** 10.1101/2021.03.21.21254074

**Authors:** Shaina L. Stacy, Hukum Chandra, Raanan Gurewitsch, LuAnn L. Brink, Linda B. Robertson, David O. Wilson, Jian-Min Yuan, Saumyadipta Pyne

## Abstract

We propose a novel, two-step method for rescaling health survey data and creating small area estimates of smoking rates using a Behavioral Risk Factor Surveillance System (BRFSS) survey administered in 2015 to participants living in Allegheny County, in the state of Pennsylvania, USA. The first step consisted of a spatial microsimulation to rescale location of survey respondents from zip codes to tracts based on census population distributions by age, sex, race, and education. The rescaling allowed us, in the second step, to utilize and select from available census tract specific ancillary data on social vulnerability for small area estimation (SAE) of local health risk using an area level version of a logistic linear mixed model. To demonstrate this new two-step algorithm, we estimated the ever-smoking rate for the census tracts of Allegheny County. The ever-smoking rate was slightly above 70% for two census tracts to the southeast of the city of Pittsburgh. Several tracts in the southern and eastern sections of Pittsburgh also had relatively high (>65%) ever-smoking rates. These small area estimates may be used in local public health efforts to target interventions and educational resources aimed at reducing cigarette smoking. Further, our new two-step methodology may be extended to small area estimation for other locations, and other health-related behaviors and outcomes.

## Introduction

In the United States (U.S.), tobacco smoking has declined considerably over the past several decades; however, an estimated 13.7% of U.S. adults still smoke cigarettes, and it is the leading cause of preventable disease, disability, and death (1). Cigarette smoking has been linked to many cardiovascular and respiratory diseases, such as chronic obstructive pulmonary disease (COPD), and is the leading risk factor for lung cancer development. Smoking cessation reduces the risk for these adverse health outcomes and can add as much as a decade to life expectancy (1). Using a combination of routinely collected health survey data and new statistical methods, we can identify neighborhoods with high smoking rates to better target smoking cessation interventions, as well as those experiencing disparities in outcomes of such programs.

National health surveys, such as the Behavioral Risk Factor Surveillance System (BRFSS) (2), are crucial tools for monitoring population trends in smoking and other high risk, health-related behaviors at the country or state level. However, local governments and other public health entities often need these population health measures at the county or subcounty level for activities such as resource allocation and targeting public health interventions, among others. National surveys alone cannot fill these needs, often due to limited coverage of small geographic areas. Further, small sample sizes of such surveys when restricted to local populations may make estimation of the variables of interest difficult and likely unreliable below the state level. To address this issue, various small area estimation techniques have been proposed to downscale national or state health survey data and generate small area estimates (SAEs) that are deemed more reliable in terms of providing insights into health conditions and health-related risk behaviors that are specific to local populations (3).

A handful of prior studies have sought to produce SAEs, such as at U.S. census tract or census block level, based on BRFSS data, including risk behaviors like smoking (4-8), health outcomes like COPD (9,10), and other factors (11,12). For example, Ortega et al. used a random effects model and census data to estimate smoking and obesity prevalence for U.S. zip codes and tracts using BRFSS data from 1991 to 2010. Overall, their SAEs were reliable, with most of the error within 2% of observed in regions with BRFSS data (5). Wang et al. applied a multilevel regression model and post-stratification method using BRFSS data to estimate the prevalence of smoking, binge drinking, and other health behaviors at a census block level, which could in turn be further aggregated to generate estimates at other geographic levels of interest (e.g., city). Although model-based estimates were consistent with direct survey estimates for many of their health indicators of interest, correlations were low for current smoking (7). Song et al. added a “nearest intersection” question to a local BRFSS survey administered in King County, Washington, to geocode data to subcounty areas, and produced smoothed estimates in cigarette smoking at census tract- and health reporting area-levels using hierarchical Bayesian models. However, the precision from their model was relatively low at the census tract level, with somewhat wide 90% confidence intervals (6).

In this study, we introduce a new two-step algorithm for survey data to rescale and generate small area estimates of the variable of interest. The term “small area” is used to describe a domain for which the sample size is not large enough to allow sufficiently precise direct survey estimation. Often indirect SAE methods depend on the availability of population level auxiliary information related to the variable of interest (3). In the first step of our algorithm, we use microsimulation for spatial “side-scaling” of the survey data from the original unit of area (e.g., at zip-code level) to a different unit of area (e.g., at census-tract level). In the process, while there might be loss of some data points due to uncertainty in their spatial assignment, the tradeoff can succeed in terms of the gain in potentially insightful auxiliary information that may be available at this re-scaled level. In the second step of our algorithm, such population level auxiliary information is used for model-based small area estimation which, in this study, is done for every census tract (or simply “tract”). We also include additional steps to decide whether to incorporate the design of the survey in our model, as well as to provide multiple model diagnostics. We then demonstrate our algorithm by computing SAEs of ever-smoking rates, leveraging a local BRFSS survey of adults residing in Allegheny County in western Pennsylvania.

## Data and Methods

The University of Pittsburgh Institutional Review Board approved this study (STUDY19040081).

### Local BRFSS Survey

The Allegheny County Health Department modeled its local BRFSS survey after the national survey, but the county raised its own funds for the survey and added many of its own questions. This county survey was administered to a random sample of adults 18 years and older who resided in Allegheny County in 2015 (13). Six percent of possible landline and 4% of cellular telephone numbers in the county were sampled, with a total of 9032 interviews secured. For the present study, we obtained these as de-identified data, with personal identifying information masked by codes. We excluded 74 survey respondents with likely erroneous ages (<18 years old) and 122 respondents with missing zip codes, leaving 8836 respondents in 105 zip-code defined areas for the spatial microsimulation (first step). Survey demographic variables (age, sex, race, and education) were re-categorized as necessary to harmonize with key census variables: sex (male or female), age (18-24, 25-34, 35-44, 45-64, ≥65 years), race (white, black, other), and education (less than high school, high school graduate, some college, and college graduate or higher).

### American Community Survey

The first step of our algorithm, spatial microsimulation, requires census population margins by demographic factors to assign survey respondents to probable tracts. The National Census takes place once every ten years (e.g., 2000, 2010, 2020); however, the American Community Survey (ACS) provides one- and five-year summary estimates for the years between the two censuses on the tract level or other geographically defined areas. The ACS is a nationwide survey that collects economic, housing, and demographic data every year. The one-year estimates have been collected over a 12-month period and are available for geographic areas with at least 65,000 people (14). We obtained 2015 tract-level population estimates from ACS to correspond to the year of our BRFSS survey.

### Social Vulnerability Data

The U.S. Centers for Disease Control and Prevention’s (CDC) Social Vulnerability Index (SVI) was originally computed to help public health officials and emergency response planners identify the most vulnerable communities that will require support during a hazardous event. The SVI ranks tracts on 15 social factors and further pools them into four summary themes: socioeconomic, household composition and disability, minority status and language, and housing type and transportation. It also provides an overall SVI (15).

### Spatial Microsimulation

Step 1 of our two-step algorithm was a microsimulation to assign survey respondents to tracts using the approach of combinatorial optimization (CO). This procedure involves the selection of an optimal combination of households from an existing survey dataset that best fit published small-area census tabulations (16). For the present analysis, a zip code-restricted CO was conducted in which the spatial microsimulation was run for each study area zip code in parallel. The assigned tracts of a pair of randomly selected respondents were swapped until an “optimal” combination of households was found to satisfy the known census population marginals. In general, CO could be computationally costly and take many iterations to converge to an optimal solution. In our case, we could restrict the swaps of individuals only among tracts which overlapped with the zip codes where the targeted individuals resided according to the survey. This allowed us to divide the CO problem into zip code-specific subproblems that were solved simultaneously, thus resulting in a computationally efficient microsimulation.

We conducted the spatial microsimulation using the simPop package in R (version 4.0.2). SimPop is an open source data synthesizer that can be used to allocate populations from larger (in our case, zip codes) to smaller geographic areas (correspondingly, tracts) (17). After the study population was initially distributed to census tracts using the simInitSpatial tool, a post-calibration procedure (calibPop) was performed to refine the distribution to tracts based on known census population marginals for age, sex, race, and education. This procedure implements CO based on simulated annealing to conduct an iterative search for a near optimal combination of households to populate the geographic areas. As this is a probabilistic step, a degree of randomness is involved in the household selection and the results will be slightly different for each run. Thus, the microsimulation was run for *N* = 100iterations for each respondent *r*. In each iteration, *r* is assigned to at most one tract within her zip code that is known from the BRFSS survey data. Further, one census table containing a population breakdown by all four demographic variables of interest was not available. We therefore repeated the microsimulation for each of the following three combinations of marginals: {age, sex, race}; {age, sex, education}; and {sex, race, education}.

Then, we spatially assign to each respondent *r* the tract which has (i) the strongest assignment among (ii) the least inconsistent of all tracts assigned to *r* by microsimulation. Let *Max*(*r, d*) and *Min*(*r, d*) be the largest and the smallest number of assignments of any tract *d* to *r* out of a total of *N =*100microsimulations of *r* for each of the three combinations of marginals as stated above. For each *r*, we sort the tracts in a sequence {*d*_(*i*)_}_*r*_ in the increasing order of *Incons*(*r, d*_*j*_) *= Max*(*r, d*_*j*_) – *Min*(*r, d*_*j*_) as long as *Incons*(*r, d*_*j*_) *< δ*. Then *r* is assigned to the first tract in the sorted sequence {*d*_(*i*)_}_*r*_ for which *Max*(*r, d*_(*i*)_) ≥ *μ*. The threshold values of *μ* and *δ* were selected as 40 and 50 based on the empirical distributions of *Max* and *Incons* to include a majority of respondents in the final assignments. If no tract met these criteria for a survey respondent, then that person was considered “unassigned” and excluded from Step 2.

### Small Area Estimation

In Step 2 of our framework, we use the rescaled microdata from Step 1 for small area estimation of ever-smoking rates for all tracts in Allegheny County. Two types of variables are used for SAE analysis. First, the variable of interest drawn from the survey, i.e., ever-smoking, which is binary at the individual level, and corresponds to whether a person had ever smoked or not. The parameter of interest was to estimate the proportion of ever smokers within each census tract (given by the 458 tracts of Allegheny County).

The second type consists of the tract-level auxiliary variables (or covariates). We used as available covariates four theme-wise summary SVI variables defined as (i) Socioeconomic: RPL_THEME1, (ii) Household Composition & Disability: RPL_THEME2, (iii) Minority Status & Language: RPL_THEME3, and (iv) Housing Type & Transportation: RPL_THEME4. These values are given as percentile ranking.

A generalized linear model between tract-specific sample (unweighted) proportions of smoking and the set of four auxiliary variables (RPL_THEME1-4) was fitted for choosing the appropriate auxiliary variables. This model was fitted using the glm function in R and specifying the family as “binomial” and the tract-specific sample size as the weight. The primary purpose was to build a good explanatory and predictive model based on the available auxiliary data. Finally, two auxiliary variables, RPL_THEME1 (Socioeconomic) and RPL_THEME3 (Minority Status & Language), which significantly explained the model, were identified for use in subsequent SAE analysis.

The final model, including the covariates RPL_THEME1 and 3, was then used to produce tract-level estimates of ever-smoking rates. The tract-specific direct survey estimates of smoking rates were defined as follows. Let *y*_*di*_ denote the variable of interest for person *i* in tract *d (d = 1, … D*). In particular, *y*_*di*_ is a binary variable that takes the value 1 if person *i* in tract *d* smokes and 0 otherwise. Here, *D* is the total number of tracts in the study population, where *D*_1_ and *D*_2_ are the number of tracts with and without sample data, respectively, such that *D*_1_ + *D*_2_ = *D*. The aim is to estimate the proportion of ever smokers, 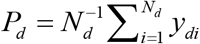, in tract *d*, where *N*_*d*_ is the population size of tract *d*. Let *w*_*di*_ be the survey weight for person *i* in tract *d*. The direct estimator (denoted by *Direct*) for *P*_*d*_ is 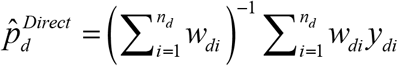, with the estimate of variance of the *Direct* estimator given by

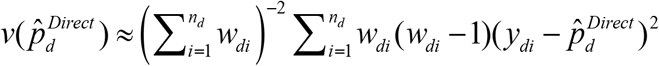, where *n_d_* is sample size for tract *d*.

In case of simple random sampling (SRS) used for survey data collection,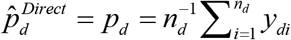 is the simple sample proportion and 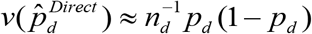, where 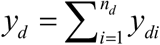 denotes the sample count in tract *d*. If the sampling design is informative, this SRS-based version of *Direct* may be biased.

Let *u*_*d*_ denote the tract-specific random effects that capture the dissimilarities between the tracts. If we ignore the sampling design, the sample count *y*_*d*_ in tract *d* can be assumed to follow a binomial distribution with parameters *n*_*d*_ and *π* _*d*_, i.e., *y*_*d*_ |*u*_*d*_ ∼ Bin(*n*_*d*_, *π* _*d*_); *d* = 1,…, *D*_1_. This leads to *E* (*y*_*d*_ |*u*_*d*_) = *n*_*d*_*π* _*d*_. Let **x**_*d*_ be the *k*-vector of covariates for tract *d* available from secondary data sources. Following previous work by study team members (16, 17), the aggregate level version of logistic linear mixed model (LLMM) linking the probability *π* _*d*_ with the covariates **x**_*d*_ is expressed as

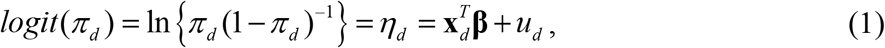

with 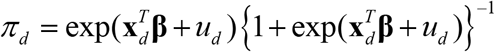. Here **β** is the *k*-vector of regression coefficients and *u_d_* is assumed to be independent and normally distributed with mean zero and variance 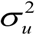. Assuming *N*_*d*_ ⋙ *n*_*d*_, an empirical plug-in predictor (EPP) of smoking proportion in tract *d* is given by

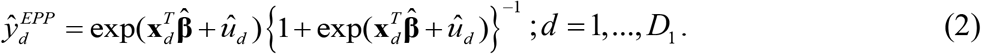

It is obvious that in order to compute the small area estimates by equation (2), the estimates of the unknown parameters **β** and 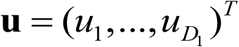 in equation (2) are obtained using an iterative procedure that combines the Penalized Quasi-Likelihood estimation of **β** and **u** with restricted maximum likelihood (REML) estimation of 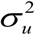 to estimate unknown parameters. For tracts with no sample data (*n*_*d*_ = 0), the synthetic type predictor of smoking proportion in tract *d* is given by

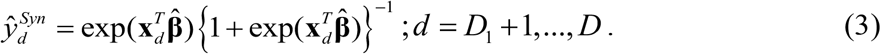

The mean squared error (MSE) estimation of small area predictor (2) and (3) is due to Chandra *et al*. (2019) (18).

### Impact of Sampling Design

In this section, we first inspect whether sampling design adopted in collecting the sample data is informative or can be ignored. The sampling design used in survey data collection must be incorporated in making the valid analytical inference about the population. For this purpose, we compute the effective sample sizes and the effective sample counts for the sample data, as described previously (18). Use of effective sample size rather than the actual sample size allows for the varying information in each area under complex sampling. Following previous work, we use the effective sample sizes in place of observed sample sizes to incorporate the sampling design (19,20).

### Diagnostic Measures

These are used for examining the assumptions of the underlying models and assessing the empirical performances of the EPP method. Generally, two types of such measures are suggested and commonly employed in SAE application; (i) the model diagnostics, and (ii) the diagnostics for the small area estimates. The main purpose of model diagnostics is to verify the distributional assumptions of the underlying small area model, i.e., how well this working model performs when it is fitted to the survey data. The other diagnostics are used to validate reliability of the model-based small area estimates.

In LLMM, equation (1), the random tract-specific effects are assumed to have a normal distribution with mean zero and fixed variance. If the model assumptions are satisfied, then the tract level random effects (or residuals) are expected to be randomly distributed and not significantly different from the regression line *y=0;* whereas, from equation (1) the area level random effects (or residuals) are defined as 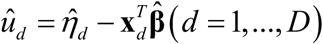. Histogram and normal probability (q-q) plot can be used to examine the normality assumption. Supplementary **Figure S1** shows the histogram (left plot), the normal probability (q-q) plot (center plot) and the distribution of the tract-level residuals (right plot). The Shapiro-Wilk test (implemented using the shapiro.test() function in R) was also used to examine the normality of the tract random effects. The value of the Shapiro-Wilk test statistic was 0.984 with 285 degrees of freedom (p-value=0.002). This indicates that the tract random effects are likely to be normally distributed. The tract level residuals appear to be randomly distributed around zero. Further, the histogram and q-q plot also provide evidence in support of the normality assumption (Supplementary **Figure S1**).

Further, a set of diagnostics described previously (21,22) are also considered for assessing validity and reliability of the tract-wise estimates generated by the EPP method. Here, we used four commonly used measures that address these requirements: a bias diagnostic, a goodness of fit test, a percent coefficient of variation diagnostic, and a 95% confidence interval diagnostic. The first two diagnostics examine the validity and last two assess the reliability or improved precision of the model-based small area estimates.

In addition, we implemented a calibration diagnostic where the model-based estimates are aggregated to higher level and compared with direct survey estimates at this level. Here0020 direct estimates DIR 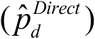 are defined as the survey weighted direct estimates. We compute bias (Bias) and average relative difference (RE) between direct 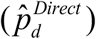 and the EPP 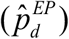 estimates as: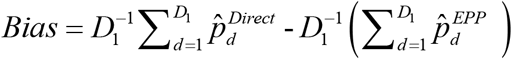 and 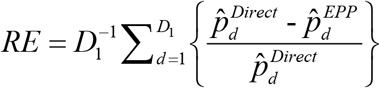 respectively.

## Results

Out of the 8836 survey respondents used for the microsimulation in Step 1, 5901 (i.e., more than two-thirds) received a final tract assignment (**Figure 1**). In general, proportions of groups by education, race, and sex across the five age categories were similar between the 2015 census and our microsimulated datasets (**Figure 2**). Out of a total of 468 Allegheny County tracts in the survey data, we had 286 tracts with samples, and the rest were out of sample. In the sample data, the sample count (i.e., number of ever-smokers in the sample) was 4517. For this study, auxiliary variables were available for 458 tracts (285 with sample data and 173 without sample data) only. Therefore, further analysis considered only 458 tracts for estimating the ever-smoking rate using SAE. At this stage, the survey data had a total sample size of 5892 respondents and sample count of 2689 (**Table 1**).

**Table 1.** Summary of Sample Size and Sample Count in Survey data.

| Characteristics | Minimum | Maximum | Average | Total |
| --- | --- | --- | --- | --- |
| Sample size | 1 | 160 | 21 | 5892 |
| Sample count (smoking incidence) | 0 | 71 | 9 | 2689 |
| Sampling fraction | 0.0028 | 0.056 | 0.0092 |  |

**Figure 1.**
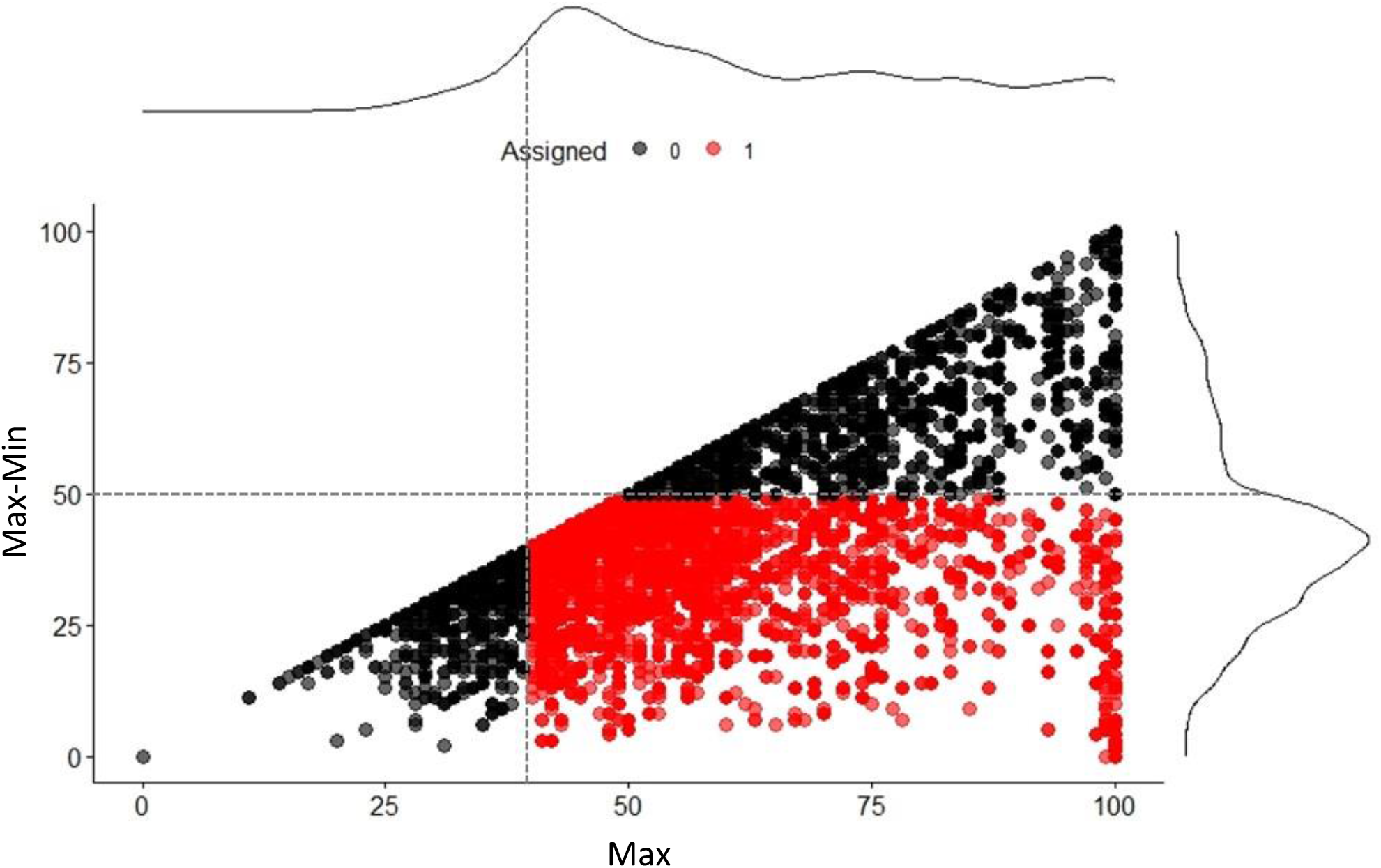
Scatterplot of *Incons* (*Max-Min*) versus *Max* values for each of *N*=8836 survey respondents due to spatial assignments in three sets of 100 microsimulations. Empirically, the dotted lines show the most inclusive thresholds at *Max* ≥40 and *Incons* <50. The resulting included assignments are shown as red dots.

**Figure 2.**
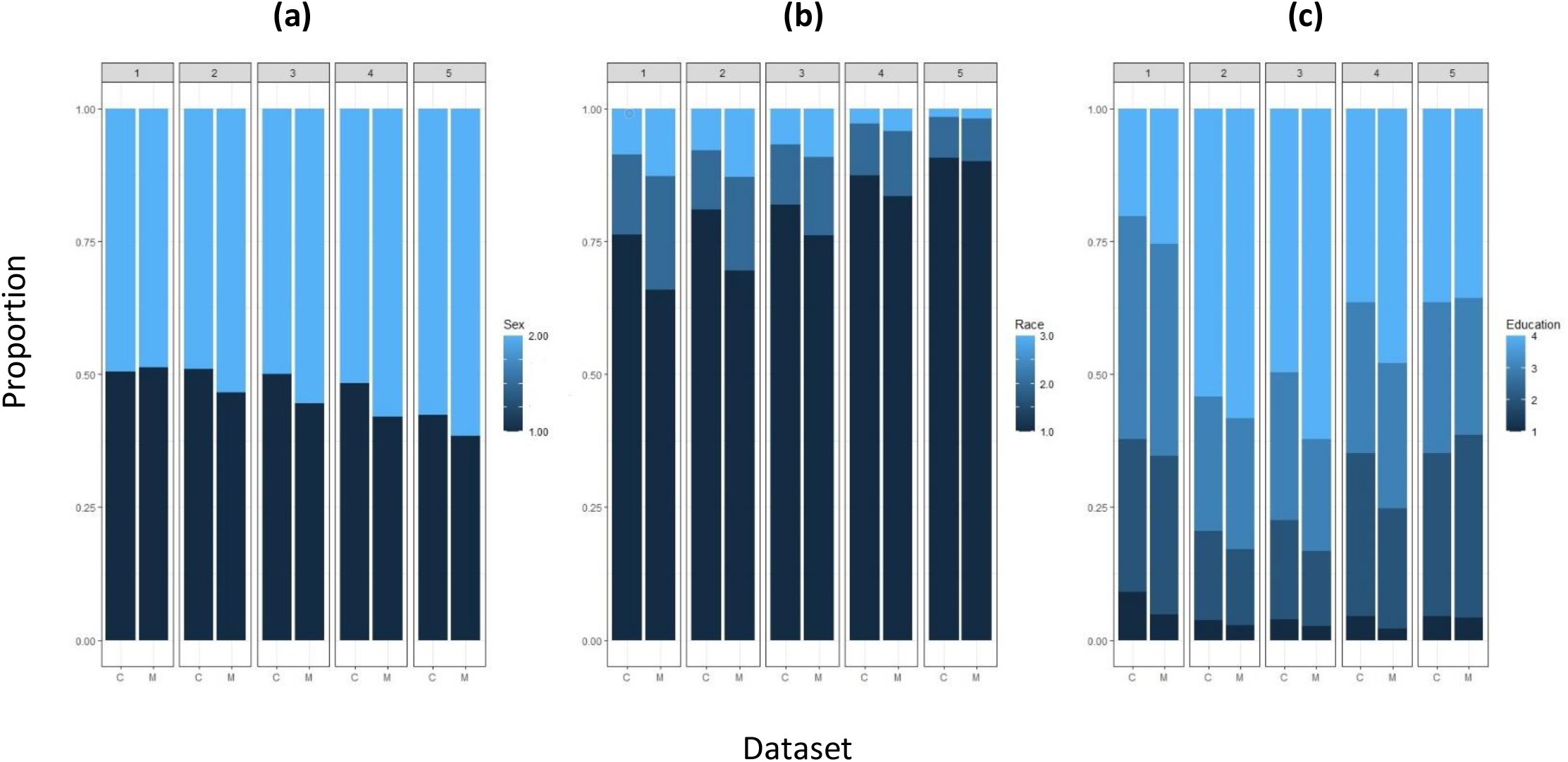
Barplots comparing the 2015 Census data (C) and Microsimulation results (M) with paired bars that show the proportions of each category of (a) sex, (b) race, and (c) education across 5 groups of increasing age.

Across tracts, the sample size ranged from one to 160 with an average of 21. The average sample count was nine per tract, with a minimum of zero and a maximum of 71. About 32% (91 out of 285) of total tracts had samples of less than five people. In the majority of tracts, the effective sample sizes are smaller than the observed sample sizes (**Figure 3**). Similarly, in most of the cases, the effective sample counts are smaller than the observed sample counts. This indicates that the sampling design is indeed informative, when compared with SRS, in such tracts. Hence, sampling weights cannot be ignored in the SAE analysis (**Figure 3, Table 1**, Supplementary **Figures S2 and S3**).

**Figure 3.**
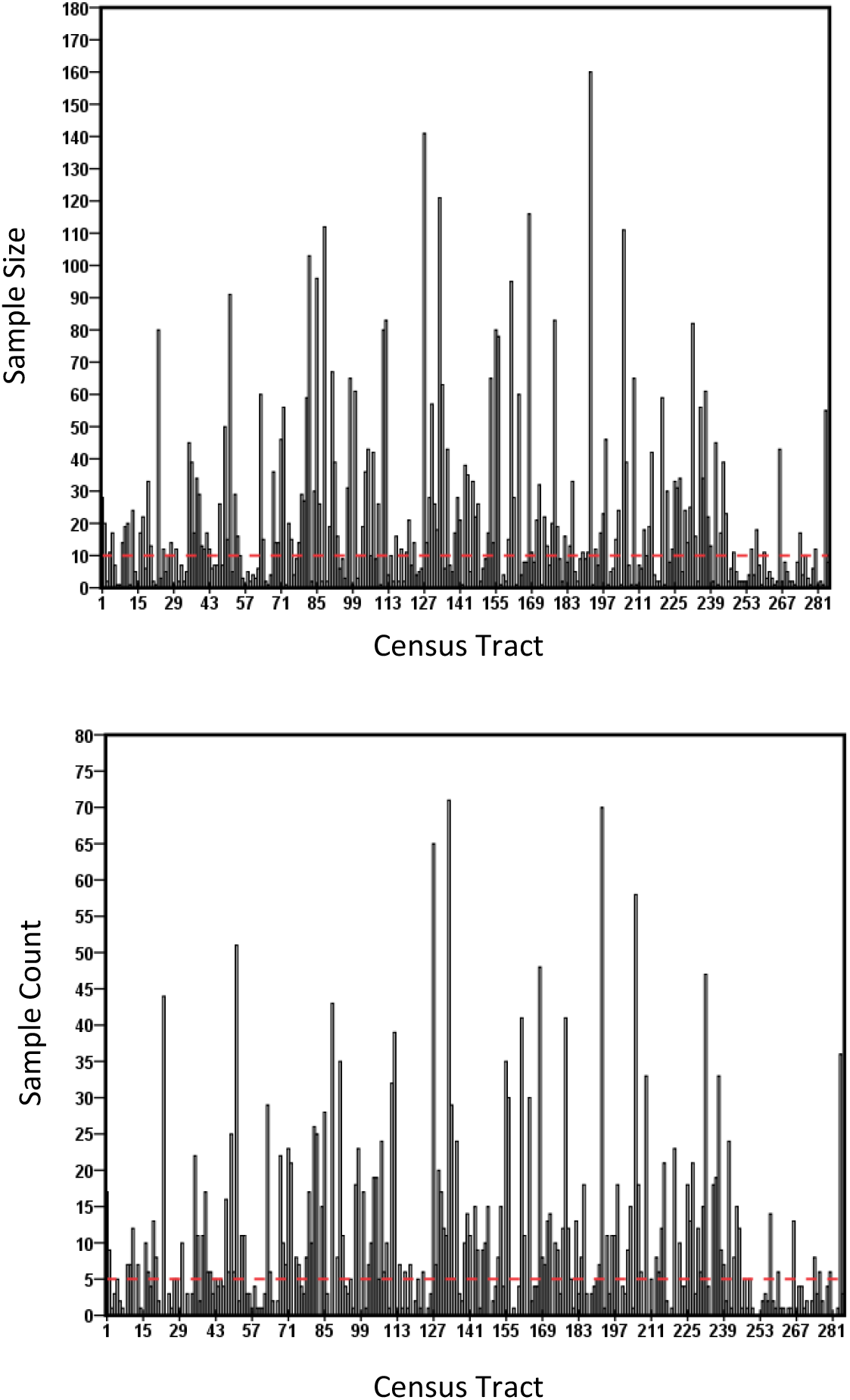
Tract-wise distribution of sample size (top) and sample count (bottom).

We fitted generalized linear models between unweighted proportions of smoking and the four SVI themes to choose the appropriate auxiliary variables. The two auxiliary variables RPL_THEME1 and RPL_THEME3 were significant predictors for the ever-smoking rate with an Akaike Information Criterion (AIC) value of 1205.5 (**Table 2**). Further, the effects of ever-smoking were positive for RPL_THEME1 and negative for RPL_THEME3. The model coefficients of RPL_THEME1 (0.82368) and RPL_THEME1 (−0.63327) were significant (*p*<0.001). The null deviance of the model was 532.35 with 284 degrees of freedom, but adding RPL_THEME1 and RPL_THEME3 in the model reduced the residual deviance to 477.55 with a loss of two degrees of freedom. RPL_THEME1 reduced the residual deviance by 31.150, while the RPL_THEME3 reduced it by 23.799, both of which were statistically significant. Using these covariates, the tract-level small area estimates, and the corresponding standard errors, were computed (available from the authors upon request).

**Table 2.** Model Parameters for the Generalised Linear Models for Smoking Rate. (_*_ *p* < 0.05; ^**^ *p* < 0.01; ^***^ *p* < 0.001)

| Parameters | Estimate | Standard Error | z value | Pr(>\|z\|) |
| --- | --- | --- | --- | --- |
| Intercept | -0.13177 | 0.06153 | -2.142 | 0.0322 * |
| RPL_THEME1 | 0.82368 | 0.11753 | 7.008 | 2.42e-12 *** |
| RPL_THEME3 | -0.63327 | 0.13015 | -4.866 | 1.14e-06 *** |
| AIC | 1205.5 |  |  |  |
| Null deviance | 532.50 with 284 df |  |  |  |
| Residual deviance | 477.55 with 282 df |  |  |  |

To validate our results, we compared our tract-level SAEs of ever-smoking rates with such estimates by a previous study (5) for the groups of years 1991-1995, 1996-2000, 2001-2005, and 2006-2010. Interestingly, the studies showed positive, significant correlations (correlation coefficients: ∼0.51, p<0.001) (**Figure 4**). However, our rate estimates ranged from 20 to 72%, whereas these prior estimates had a narrower spread (∼10-40%). In our analysis, the tracts with the highest estimated ever-smoking rate, slightly over 70%, were located southeast of the city of Pittsburgh. Other tracts with relatively high rates (>65%) were located within neighborhoods in the southern (Hazelwood, Arlington, Carrick) and eastern (East Hills) sections of Pittsburgh. There was also a cluster of tracts with relatively high rates to the west of Pittsburgh (**Figure 5a**). As expected, the standard errors of SAE are higher in non-sample tracts (**Figure 5b**). Distributions were similar between tracts in the city of Pittsburgh versus outside of Pittsburgh, although the SAEs for non-city tracts had slightly more spread (Supplementary **Figure S4**).

**Figure 4.**
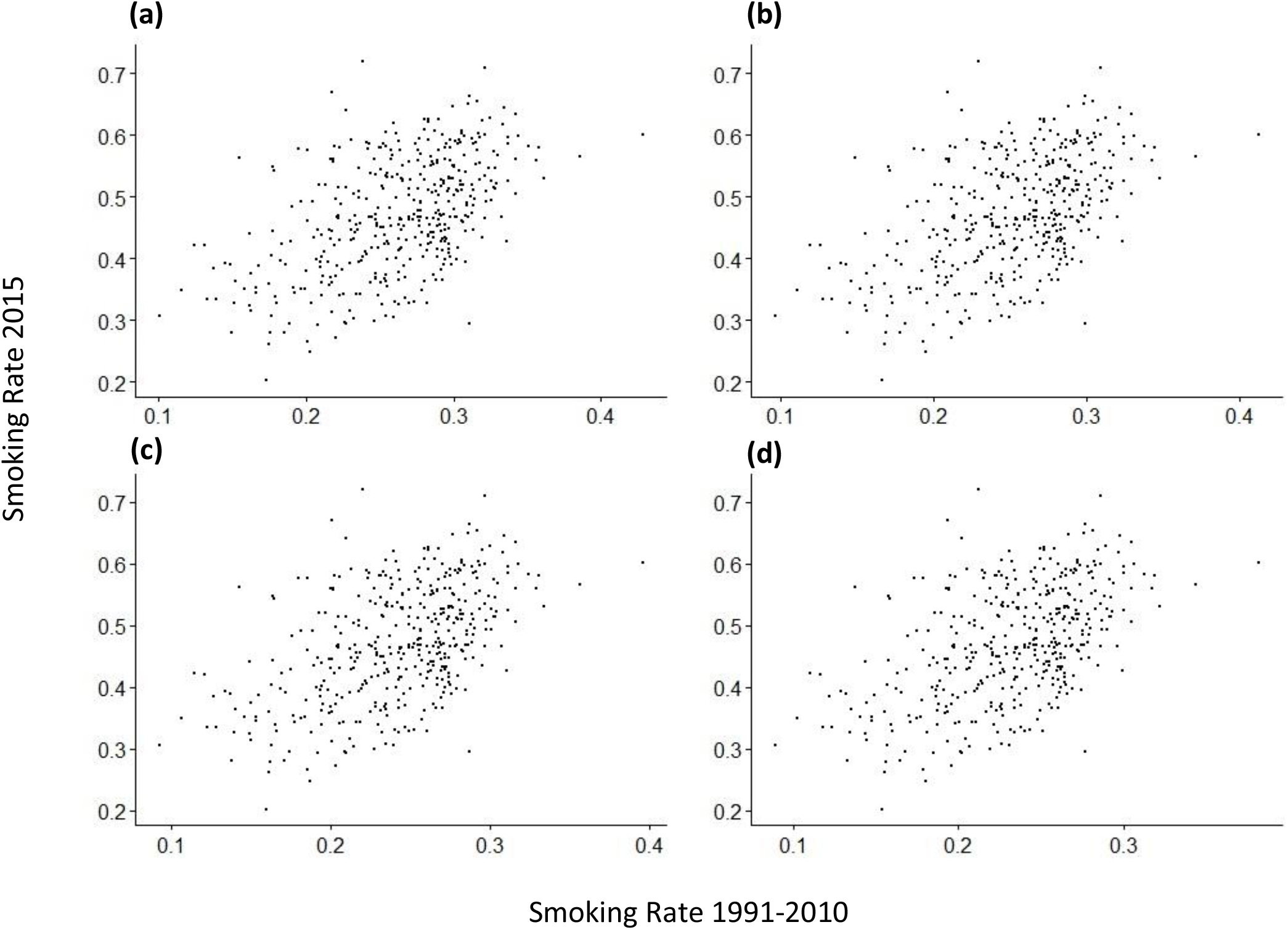
Scatterplots of SAEs of smoking rates calculated for 2015 (y-axis) in this study versus SAEs due to Oretega et al. for the years: (a) 1991-1995, (b) 1996-2000, (c) 2001-2005, and (d) 2006-2010.

**Figure 5.**
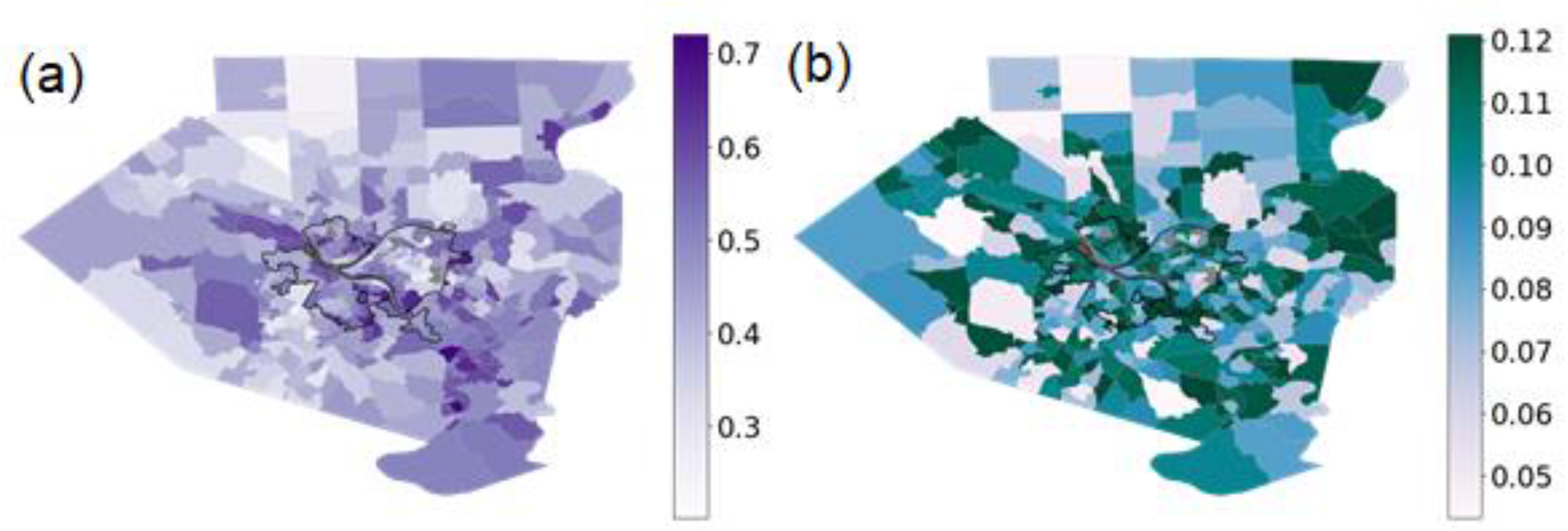
The maps show (a) the small area estimates and (b) standard errors (SE) of smoking rates in each tract of Allegheny County. The bold, black outline delineates the city of Pittsburgh.

Finally, our small-area, ever-smoking rate estimates may be considered in the context of lung cancer, a major health effect of cigarette smoking, and concomitant exposures. Here, we take the example of radon as it is considered the primary risk factor for lung cancer among non-smokers and may have a synergistic effect with smoking to increase lung cancer risk (23). There were a handful of tracts with high ever-smoking rates that were also among the highest for age-adjusted incidence rates of lung cancer calculated for the period 2011-2017 (Supplementary **Figure S5**). Some tracts appeared to have relatively higher smoking rates, average radon levels, and lung cancer incidence. Notably, some tracts (e.g., tract 5128) with high lung cancer incidence (150 per 100,000 people) had relatively lower smoking rates (0.36) but higher proportions (0.63) of household radon measurements that exceeded the U.S. Environmental Protection Agency (E.P.A.) action level of 4 pCi/L. Such observations could lead to further investigation of exposures at local levels.

## Discussion

Aggregation of data at different spatial scales can lead to scale-specific statistical bias in the form of modifiable areal unit problem (MAUP) (24). To avoid MAUP, researchers may draw inferences at a scale that best suits the particular issue of interest such as for administrative decision-making at subcounty levels, say, for optimal resource allocation. In addition, for technical reasons, they may consider certain levels to be less suitable (e.g., zip codes can change over time) or more suitable (e.g., the availability of census data for census tracts). Often, researchers address such practical concerns as “data transformation” using ad hoc aspatial approaches. The objective of our study was to provide a methodical approach to locally re-assign the microdata to the desired spatial scale, especially one that will then allow the use of local covariates to guide the inference.

In this study, we have constructed a novel, two-step algorithm for rescaling health survey data and modeling the rate of small area-level, health-related outcomes or behaviors. We used Allegheny County as a case study to demonstrate our proposed methodology and estimated ever-smoking rates at the tract level. Health surveys, including the BRFSS and others, often do not provide spatial resolution below the state or county level. The local BRFSS survey administered in Allegheny County did collect zip code of residence, but without tract assignments, linkage with informative, ancillary data sources, such as the SVI, is difficult. Our microsimulation step allowed us to distribute survey respondents to tracts within the study area in a way that reflected the known sociodemographic composition of the tracts. While not every survey respondent may meet the criteria to receive a final tract assignment, we gained in spatial resolution in terms of those that were assigned during the rescaling process.

According to the most recent Surgeon General’s report, 13.7% of U.S. adults smoke (1). Although the adult smoking rate in Allegheny County decreased from 23% in 2009-2010 to 19% in 2017 (25), this still exceeds the national rate. Racial disparities also persist in the county, both for smoking and smoking-related health outcomes. African Americans are both more likely to smoke (30% compared to 17% of whites) and have rates of lung cancer 15-30% higher than whites (13). The small area estimates of smoking rates demonstrated in this study, and its rigorous use of tract-specific (socioeconomic and minority & language based) vulnerability covariates in the estimation, could inform local smoking cessation interventions to further decrease smoking rates in the county, particularly for high-risk groups. In addition, lower socioeconomic communities face greater burdens of environmental pollution (26), further compounding their risks for cancer and other diseases. The smoking rates estimated in this study would be useful in future studies of respiratory diseases, including lung cancer, and concomitant environmental assaults. For example, an estimated 46% of tracts in Allegheny County have radon concentrations exceeding the U.S. E.P.A’s threshold of 4 pCi/L. Radon is thought to be the primary contributor to lung cancer risk among never smokers and may also act synergistically with tobacco smoking to increase lung cancer risk in smokers (23). When examining the distribution of age-adjusted lung cancer rates against radon levels and ever-smoking rates in Allegheny County, there are several tracts that are relatively high for all three variables (Supplementary **Figure S5**).

Our new two-step algorithm, combining a microsimulation step with small area estimation of tract-level smoking rate, is a major strength of our study. Further, the use of a local BRFSS survey, which contained zip codes of residences and individual-level demographic information, provided an informative dataset for rescaling respondents to tracts based on age, sex, education, and race. We applied a logistic linear mixed model with tract-specific social vulnerability covariates, and used effective sample size and effective sample count to account for the sampling design used in the survey. The two-step methodology outlined here is flexible for future application to other health surveys and outcomes.

Past applications of SAE on BRFSS data, e.g., Zhang et al. (2014), were based on fitting a unit level logistic linear mixed model to BRFSS data and then drawing 1000 random samples from their estimated conditional distributions using the fitted model parameters, and thus, generating a sample of 1000 small area estimates for each small area defined in the study (9). The efficacy of the generated small area estimates is therefore highly dependent upon the fitted model. The SAE method under an area level, logistic linear mixed model applied in this paper is a widely used approach if the model covariates (e.g., census variables) are only available in aggregate form. This approach has a simple and closed form expression and, therefore, practitioners of small area methodology as well as national statistical agencies (e.g., Office for National Statistics, Australian Bureau of Statistics, etc.) often prefer it.

Yet, our study has multiple limitations. The spatial re-scaling in Step 1 to gain in terms of the ability to include insightful covariates has a potential cost in terms of some loss of power. The CO method used is probabilistic, and thus, a degree of randomness is involved in the spatial assignment of respondents into tracts (16). In Step 2, as one would expect, standard errors were higher among non-sample compared to sample tracts. Caution should be used in interpreting the SAE results in these non-sample tracts. We do not have reliable, direct-estimate data to validate our SAE census tract results, although they correlate significantly with those from past studies. Finally, while these tract-level estimates may be used to target smoking cessation interventions or help identify high-risk communities for smoking and related health outcomes, they cannot be used to draw inferences about smoking habits of specific individuals residing in the small areas.

In conclusion, we proposed a two-step method for rescaling survey data to more granular geographic levels for which ancillary data may be available to produce locally relevant estimates for health-related risk behaviors at these levels. We used smoking rates in Allegheny County both as a case study to demonstrate this algorithm as well as to create tract-level estimates that may be used in local public health interventions or additional studies. Future work could leverage the methods described here for other health surveys, locations, diseases, and health-related behaviors.

## Data Availability

Supplemental data are available from the authors upon request.

## Supplemental Material

The Supplemental Material includes Figures S1-S5.

## Abbreviations

ACS: American Community Survey
AIC: Akaike Information Criterion
BRFSS: Behavioral Risk Factor Surveillance System
CO: Combinatorial Optimization
COPD: Chronic Obstructive Pulmonary Disease
EPP: Empirical Plug-In Predictor
LLMM: Logistic Linear Mixed Model
MAUP: Modifiable Areal Unit Problem
MSE: Mean Squared Error
SVI: Social Vulnerability Index
U.S. E.P.A.: United States Environmental Protection Agency

## Acknowledgements

This work was supported by grant P30CA047904 from the UPMC Hillman Developmental Funding Program. The authors declare that they have no conflicts of interest.

