## Supplemental file for "Rescaling and Small Area Estimation of Health Survey Data as applied to Smoking Rates in Allegheny County, Pennsylvania"

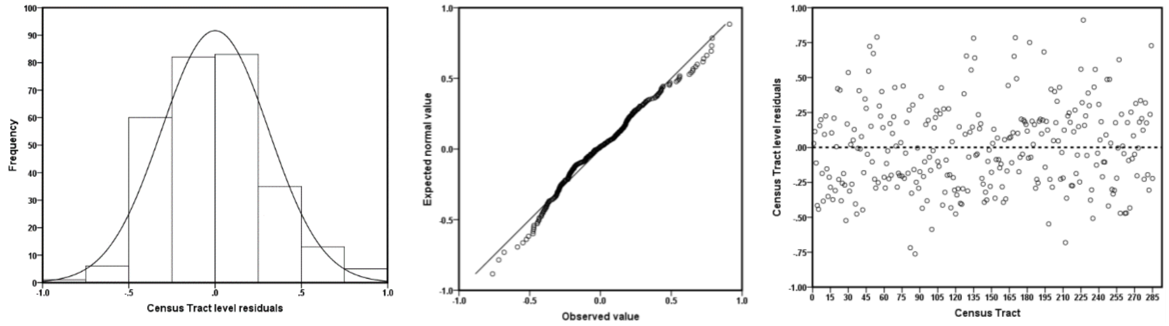


**Figure S1.** Histograms (left plot), normal q-q plots (center plot) and distributions of the tract level residuals (right plot).

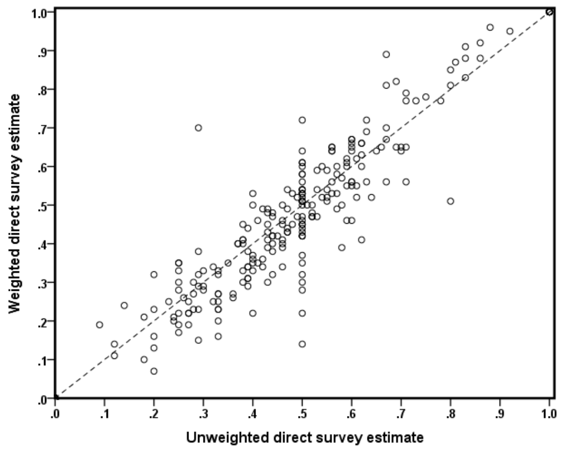


**Figure S2.** Tract-wise survey weighted vs. unweighted direct estimates of smoking rates.

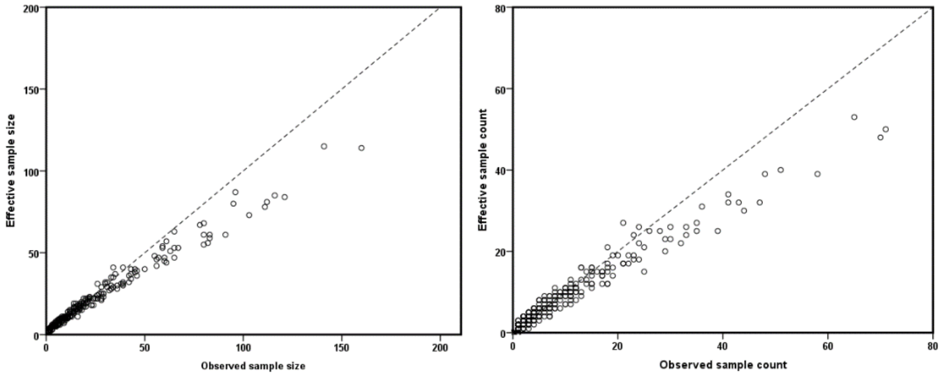


**Figure S3.** Tract-wise effective sample size vs. observed sample size (left) and effective sample count vs. observed sample count (right).

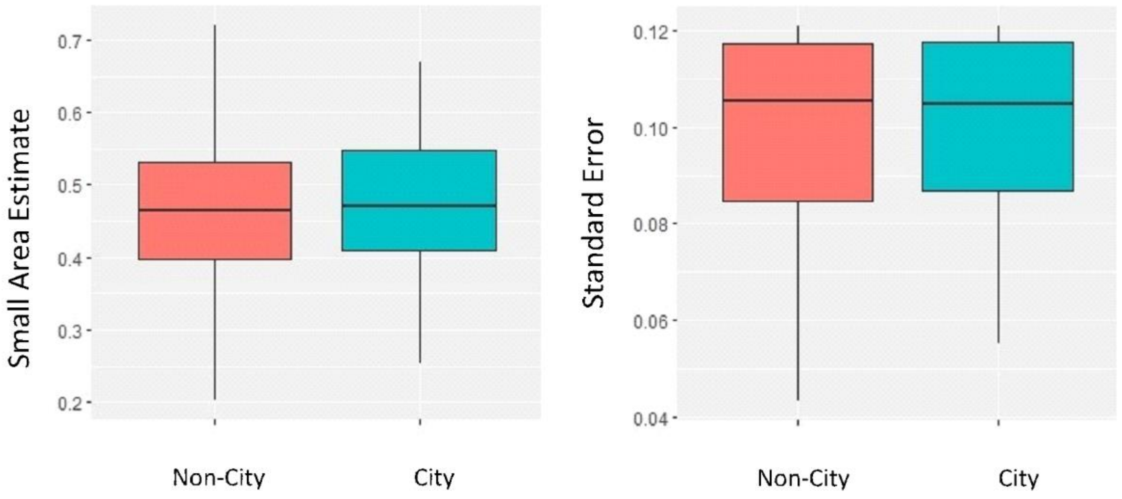


**Figure S4.** Box plots comparing small area estimates of smoking rates and standard errors between the city of Pittsburgh and non-Pittsburgh tracts.

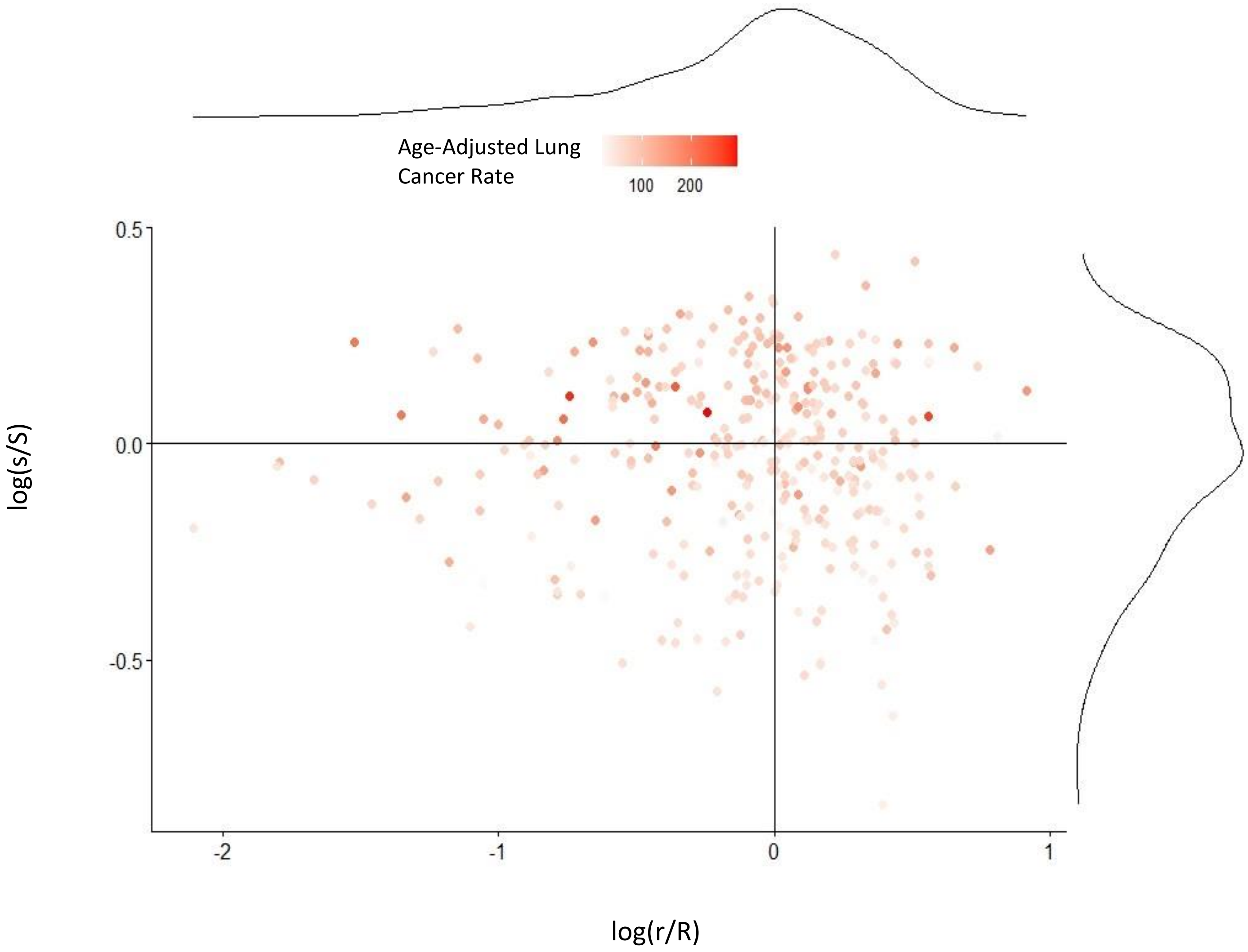


**Figure S5.** The log(s/S) versus log(r/R) by tract and time period, where s and r denote the smoking rate and the proportion of household samples with high radon levels (>4 pCi/L) for a given tract, and S and R are the corresponding median values. The shading represents age-adjusted lung cancer incidence rates for the years 2011-2017.
